# SARS-CoV-2 infection risk among unvaccinated is negatively associated with community-level vaccination rates

**DOI:** 10.1101/2021.03.26.21254394

**Authors:** Oren Milman, Idan Yelin, Noga Aharony, Rachel Katz, Esma Herzel, Amir Ben-Tov, Jacob Kuint, Sivan Gazit, Gabriel Chodick, Tal Patalon, Roy Kishony

## Abstract

Mass vaccination has the potential to curb the current COVID-19 pandemic by protecting vaccinees from the disease and possibly lowering the chance of transmission to unvaccinated individuals. The high effectiveness of the widely-administered BNT162b vaccine in preventing not only the disease but also infection suggests a potential for a population-level effect, critical for disease eradication. However, this putative effect is difficult to observe, especially in light of highly fluctuating spatio-temporal epidemic dynamics. Here, analyzing vaccination records and test results collected during a rapid vaccine rollout for a large population from 223 geographically defined communities, we find that the rates of vaccination in each community are highly correlated with a later decline in infections among a cohort of under 16 years old which are unvaccinated. These results provide observational evidence that vaccination not only protects individual vaccinees but also provides cross-protection to unvaccinated individuals in the community.

---

The recently authorized Pfizer-BioNTech COVID-19 BNT162b vaccine is highly effective at preventing disease and infection at the individual level, as demonstrated in the clinical trial, as well as in real-world vaccination campaigns^1–4^. Moreover, among vaccinees infected with SARS-CoV-2, a lower viral load was observed^5^. Reduced infection and viral load suggest reduced transmission. However, vaccination could, in principle, also increase transmission due to behavioral effects; as vaccinees may not quarantine after contacting a COVID-19 patient, or be less mindful of social distancing measures^6^. It is therefore unclear whether, overall, vaccination reduces transmission, thereby conferring protection to those who are unvaccinated or cannot currently be vaccinated, for example individuals below 16 years old, or individuals with a poor immune response^4,7–9^. However, since even in absence of vaccination the basic reproduction number varies with sociobehavioral and environmental factors, and global disease rates represent both vaccinated and unvaccinated individuals, it has proven challenging to determine the effect of vaccination on community-level SARS-CoV-2 transmission^10–12^.

The rapid vaccine rollout in Israel, initiated on December 19, 2020 and covering almost 50% of the population within 9 weeks, presents a unique opportunity to test these questions using real-world data. Capitalizing on differences in vaccination rates among geographically distinct communities, and on the availability of an unvaccinated bystander cohort of individuals below 16 for whom the vaccine is not authorized, we asked whether and to what extent the fraction of patients vaccinated in each community affects the risk of infection in an unvaccinated cohort of under 16 years old within this same community.

We focused our analysis on the vaccination rates and test results of 223 distinct communities which experienced a similar epidemic pattern prior to vaccination. We retrieved vaccination records and test results of members of Maccabi Healthcare Services, Israel’s second largest healthcare maintenance organization. By January 30th 2021, 644,609 out of 1.95 million of MHS members, residing in 279 distinct communities throughout Israel, were vaccinated with at least a single vaccine dose. To minimize a potential effect of intrinsic differences between communities and isolate the putative protective effect of vaccination rates on the unvaccinated population, we first clustered communities based on their temporal patterns of COVID-19 infection rate prior to vaccination onset (Supplementary Fig. 1; see Methods, *Clustering communities*). We identified two clearly distinct temporal patterns of infection: a large cluster of 223 communities accounting for 79.9% of communities and 69% of MHS members, and a smaller cluster of 56 communities. These clusters differed in their pre-vaccination community-level positive test rates, with a median positive rate of 3.6% (IQR: 1.9%) and 14.2% (IQR: 5.5%) for the large and small clusters, respectively. As both vaccination and natural infection may render individuals immunized, thereby possibly conferring protection on unvaccinated individuals, high infection rates may mask the effect of vaccination-induced immunity. We therefore chose to focus on the community cluster with lower positive rates, which will allow us to identify vaccination effects while minimizing the potential confounding effect of natural immunization.

To control for the spatio-temporally dynamic nature of the epidemic, we focused on relative changes in positive test rates within each community at fixed time intervals, thus accounting for global temporal patterns as well as for intrinsic differences among communities. First, for each community, the mean cumulative fraction of patients inoculated with the first dose of the vaccine was calculated at three distinct intervals for individuals 16-50 years old, which presumably represent the population likely to interact with the bystander unvaccinated cohort of under 16 years old (Fig. 1a; *V*_1_, *V*_2_, and *V*_3_). Then, for each such vaccination time interval, a corresponding time-shifted testing interval was defined with a delay of 35 days, to allow for the putative immunization of vaccinees to take effect (Fig. 1a,b). For these three testing intervals, the positive rate among the unvaccinated bystander cohort was calculated (Fig. 1b, *P*_1_, *P*_2_, and *P*_3_). Comparing periods 1 and 2, and periods 2 and 3, we then focused on the relationship between the change in positive rate of the unvaccinated cohort (*P*_2_/*P*_1_, *P*_3_/*P*_2_) and the change in vaccination (*V*_2_-*V*_1_, *V*_3_-*V*_2_; Fig. 1c,d).

**Figure 1:**
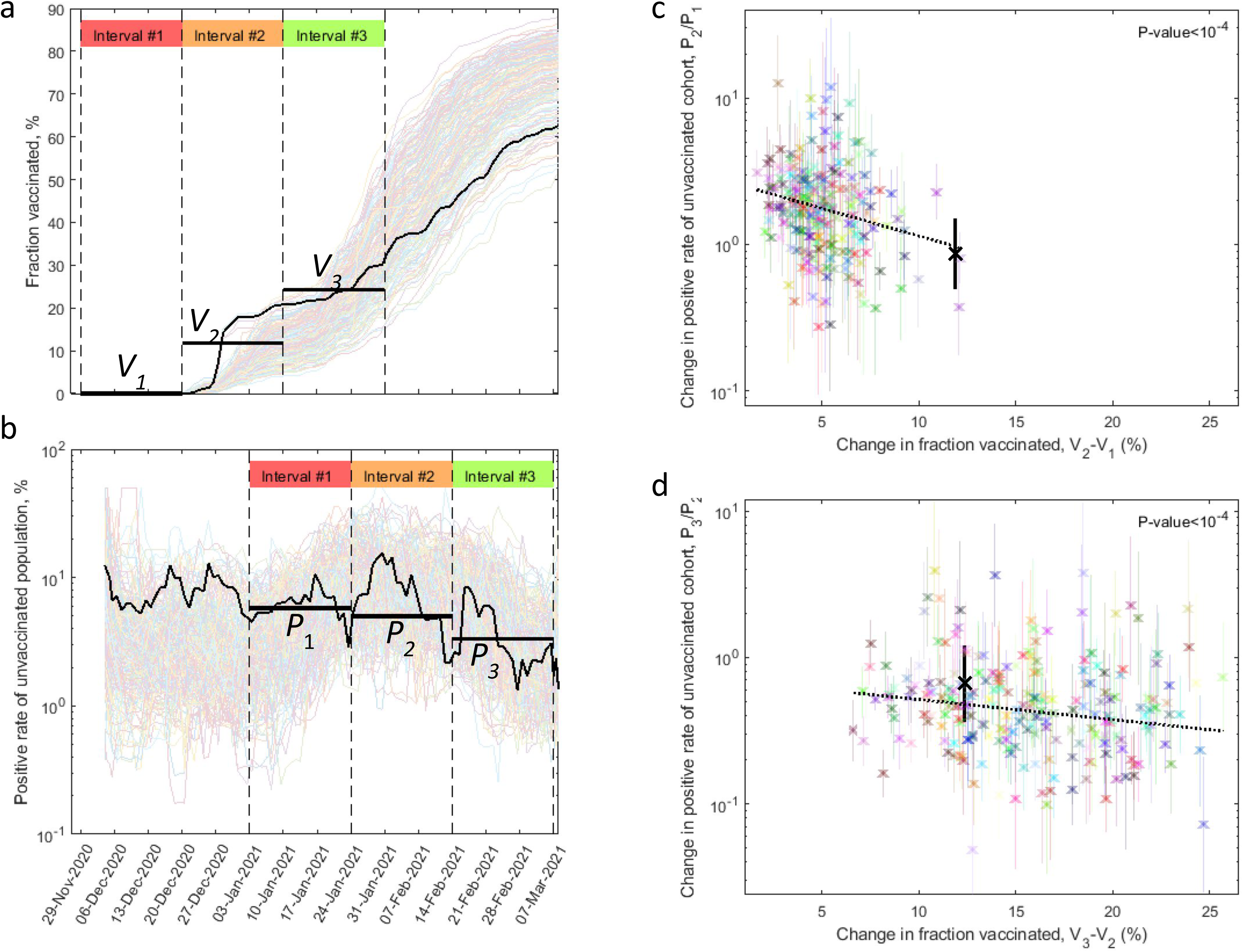
Increased fraction of vaccination in a community is associated with a later reduction in infection rate of the unvaccinated cohort in the same community. (**a**,**b**) The accumulated fraction vaccinated among 16-50 years old (a) and the positive test rate among the 0-16 bystander unvaccinated cohort (b, 7-day moving average) is shown for each community as a function of time. The three intervals for measuring vaccination (a) and their corresponding time-shifted intervals for measuring positive rates (b) are indicated on top of each panel (red, orange, green). The black trajectory highlights an example community, indicating its mean vaccination fraction in each of the intervals (*V*_1_, *V*_2_, *V*_3_), and the mean positive rates of the unvaccinated cohort in the later corresponding intervals (*P*_1_, *P*_2_, *P*_3_; see Methods, *Vaccination and testing intervals*). (**c**,**d**) For each community, the change in positive rate between consecutive time intervals (c, *P*_2_/*P*_1_; d, *P*_3_/*P*_2_) is shown as a function of the change in vaccinated fraction in the corresponding time intervals (c, *V*_2_-*V*_1_; d, *V*_3_-*V*_2_). Dash line shows linear fit (c, P-value<10^−4^; d, P-value<10^−4^; see Methods, *Correlation analysis*). The community highlighted in panels a,b is indicated (black).

The risk of infection in the unvaccinated cohort decreased in proportion to the rate of vaccination in each community. A linear fit showed a strong negative association between the log odds ratio of the positive rates and the increase in vaccination fraction both when comparing intervals 1 and 2 (Fig 1c, *log*(*P*_2_/*P*_1_)=*a*(*V*_2_-*V*_1_)+*b, a*=−8.7 [95% CI: −12.3 to −5.1], P-value<10^−4^) and when comparing intervals 2 and 3 (Fig 1d, log(*P*_3_/*P*_2_)=a(*V*_3_-*V*_2_)+*b, a*=−3.2 [CI: −4.8 to −1.5], P-value<10^−4^; see Methods, *Correlation analysis*). Moreover, a strong negative correlation was also observed when considering the absolute vaccination fraction at each interval (*log*(*P*_2_/*P*_1_)=*a*·*V*_2_+*b, a*=−8.7 [CI: −12.3 to −5.1], P-value<10^−4^; *log*(*P*_3_/*P*_2_)=*a*·*V*_3_+*b, a*=−2.2 [CI: −3.4 to −0.9], P-value<10^−3^). The correlations remained strong when the upper age limit for measuring vaccination rate was lowered to 40 (P-value<10^−4^ for both time interval comparisons) or increased to 60 (P-value<10^−4^ for intervals 1 and 2, and P-value<10^−3^ for intervals 2 and 3), but became insignificant when the age limit was increased beyond 70, possibly reflecting less frequent contacts between the vaccinated and unvaccinated cohorts. Taken together, our analysis shows strong negative association between vaccination of adults in the community and a later decrease in infection of the bystander young cohort.

Our study has several important limitations. First, beyond vaccine-based immunity, infections of unvaccinated could also be affected by naturally acquired immunity. While we minimize this confounding factor by focusing on communities with low cumulative prior infections, future studies may more directly control for this effect, for example by including seroprevalence data. Second, individual behavior and public policy guidelines may correlate with vaccination rates, and affect the infection potential of the unvaccinated group. Third, the proportion between the measured positive test rate and the actual infection rate could be different in each community and may also vary with time. Using the logarithmic derivatives [*log*(*P*_*2*_/*P*_*1*_), or *log*(*P*_*3*_/*P*_*2*_)], our analysis accounts for the possibility of different proportions between the infection rate and the positive rate in each community as well as for the possibility that this proportion changes uniformly with time, but it cannot account for the possibility that these proportions change over time in different ways for different communities. Finally, our study population is limited to MHS members, only partially representing the overall population for each community.

In this study, we identified a strong negative association between vaccination rate at the community level and the risk of infection for unvaccinated members of the community. We find that higher vaccination rates were associated with a later lower infection rate among the unvaccinated cohort. While the observed vaccine-associated protection of unvaccinated is encouraging, further studies are required to understand whether and how it might support the prospect of herd immunity and disease eradication.

## Methods

### Data collection

Anonymized electronic health records were retrieved for the study period: July 5th 2020 - March 9th 2021. These records include: (a) Patient demographics, indicating for each MHS member: a random ID used to link records, year of birth, and a coded geographical location of residence. For anonymization purposes, location of residence was given as random codes of (1) city/town, and (2) Geographical Statistical Area (GSA, The country is divided by the National Bureau of Statistics to GSAs corresponding to areas or neighborhoods). (b) Test results, indicating for any SARS-CoV-2 RT-qPCR test performed for MHS members: the patient random ID, the sample date, and an indication of positive and negative results (total 3,454,388 tests, with 192,643 positive results). (c) Vaccination, indicating patient by random ID and date of inoculation with the first dose of the BNT162b2 mRNA COVID-19 (1.37 million vaccinees).

### Vaccination and testing intervals

We defined 3 consecutive three-weeks testing intervals between January 3rd 2021 and March 6th. For each of these three testing intervals and each community, the positive test rate among unvaccinated individuals below 16 years old was calculated (*P*_1_, *P*_2_ and *P*_3_), and their standard errors were evaluated assuming poisson statistics. MHS members were excluded from this calculation if they were tested more than 20 times (to avoid patients participating in extensive surveys). In addition, for each MHS member, any test following a positive test was excluded from this calculation. To account for the expected time post first dose for a vaccinee to acquire immunity and to potentially prevent further infections, each three-week testing interval was matched with a corresponding three-week vaccination interval time-shifted backwards by 35 days. For each vaccination interval and each community, the mean of the cumulative fraction of individuals aged 16-50 vaccinated with the first vaccine dose (out of all MHS members aged 16-50 in the community) was calculated (*V*_1_, *V*_2_ and *V*_3_).

### Defining communities

Communities were defined as members residing in the same GSA geographical code. For each city code, all GSAs with less than 2000 members were joined into a single community. Communities were excluded if (1) the total number of tests for any of the 12 two-weeks time bins of the pre-vaccination period was smaller than 30 (too noisy for clustering, see below), (2) the number of tests for any seven days time window following vaccination was smaller than 2 (too noisy for positive rate over time, Figure 1b), (3) the number of tests for any of the three testing intervals was smaller than 50 (too noisy for correlation analysis, Figure 1c,d), or (4) the number of positive tests for any of the three testing intervals was zero (would cause a division by zero or a log of zero when calculating Y values for the scatter plots, Figure 1c,d). This analysis resulted in 279 communities. Members not residing in one of these eligible communities were excluded from our analysis.

### Clustering communities

Positive test rates were calculated for each community, for the pre-vaccination period between July 5th and December 19th in two-weeks time bins, resulting in 279 vectors (communities) of length 12 (time). Community vectors were then clustered using MATLAB’s *pdist* and *linkage* built-in functions. The clustering result was used to group the communities into two clusters, containing 223 and 56 communities.

### Correlation analysis

For each two consecutive three-weeks intervals (intervals 1 and 2, and intervals 2 and 3), a linear fit was performed on the log ratio of the positive rate and the change in vaccination using MATLAB’s *glmfit* built-in function (Y=*log*(*P*_2_/*P*_1_) and X=*V*_2_-*V*_1_ for intervals 1 and 2; Y=*log*(*P*_3_/*P*_2_) and X=*V*_3_-*V*_2_ for intervals 2 and 3). For each of the two linear fits, the slope P-value was calculated as the fraction of smaller or equal slopes calculated in 10,000 X-Y permutation bootstrapping simulations.

### Ethics committee approval

The study protocol was approved by the ethics committee of Maccabi Healthcare Services, Tel-Aviv, Israel. The IRB includes an exempt from informed consent.

## Data Availability

Due to patient privacy concerns, data is only available through a remote server, pending MTA.

## Figure Captions

**Supplementary Figure 1:**
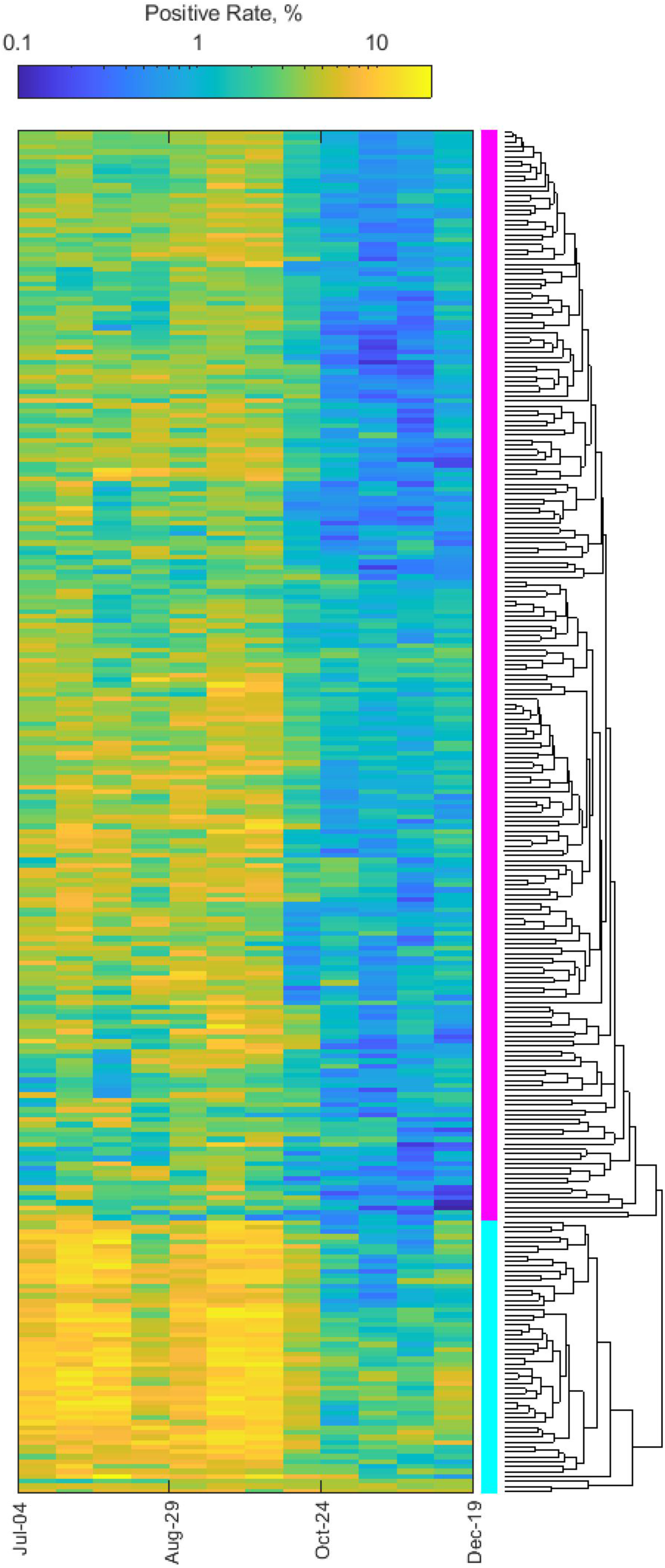
Communities are clustered based on similarity in their pre-vaccination disease dynamics. The positive rate for each community (color map) is shown as a function of time prior to the last day before vaccine rollout (December 19th, 2020). Data is binned into two-weeks periods. Cities are clustered based on euclidean distance among their positive rates, yielding two distinct clusters (dendrogram, right). The top (magenta) cluster, of 223 communities with relatively low infection rate, was chosen for analysis of vaccine effects.

